# Neighborhood-Level Public Facilities and COVID-19 Transmission: A Nationwide Geospatial Study In China

**DOI:** 10.1101/2020.08.25.20181362

**Authors:** Xurui Jin, Yu Leng, Enying Gong, Shangzhi Xiong, Yao Yao, Rajesh Vedanthan, Chenkai Wu, Lijing L. Yan

## Abstract

Individual-level studies on the coronavirus disease 2019 (COVID-19) have proliferated; however, research on neighborhood-level factors associated with COVID-19 is limited. We gathered the geographic data of all publically released COVID-19 cases in China and used a case-control (1:4 ratio) design to investigate the association between having COVID-19 cases in a neighborhood and number and types of public facilities nearby. Having more restaurants, shopping centers, hotels, living facilities, recreational facilities, public transits, educational institutions, and health service facilities was associated with significantly higher odds of having COVID-19 cases in a neighborhood. The associations for restaurants, hotels, reactional and education facilities were more pronounced in cities with fewer than six million people than those in larger cities. Our results have implications for designing targeted prevention strategies at the neighborhood level to reduce the burden of COVID-19.

---

Coronavirus disease (COVID-19) caused by the SARS-CoV-2 virus has claimed over ten million cases reported in 185 countries or regions by July 30, 2020 (1). Through massive societal efforts including a city-wide lock-down in Wuhan and nation-wide social distancing, the outbreak in China from January to April, 2020 has been contained (2). Studies on COVID-19 have proliferated since early 2020, with the majority focusing on individual-level risk factors for COVID-19 transmission and clinical management of COVID-19 patients. (*3-6*). However, neighborhood-level factors have been relatively understudied, despite contributing to infectious risk and being potential targets for disease prevention and management. For instance, prior studies have demonstrated that neighborhood-level public facilities, such as surrounding gyms and restaurant, are associated with infectious disease transmission, particularly in diseases transmitted by contact, aerosols, or droplets (*7-10*). However, little is known about the potential role of these types of public facilities in COVID-19 transmission.

Since the early stage of the COVID-19 pandemic, Chinese local governments and Centers for Disease Control and Prevention (CDC) regularly released online the names of neighborhoods with confirmed cases of COVID-19. This was done primarily to increase outbreak transparency and risk awareness for nearby residents, but it also provided a valuable data source for neighborhood-level geospatial studies. One of the defining features of neighborhoods is the number of public facilities surrounding the living areas. These facilities include gyms, restaurants, and parks, where residents, particularly older adults, spend most of their time when not at home (*11, 12*), and are thus highly relevant for potential susceptibility to COVID-19 (13).

In the present study, we aimed to examine the associations of the numbers and types of neighborhood-level public facilities with COVID-19 risk. Such information can be important for urban planners, policymakers, and public health professionals to try and reduce neighborhood transmission of COVID-19. Details of the study methods are given in supplementary materials [Section 1]. A condensed version is provided here.

Briefly, we collected latitude and longitude coordinates (geocodes) of neighborhoods with publicly released COVID-19 cases from January 18^th^ to April 30, 2020 through two Application Programming Interfaces. We also extracted all points-of-interest (POIs) data on the names, locations, and types of various public facilities from Autonavi (*Gaode*), a Chinese desktop web mapping service application. Consistent with previous studies on POIs and the classification in *Gaode* map (*17*), we classified the POI data into eight types including restaurants, shopping centers, hotels, living facilities, recreational facilities, public transits, educational institutions, and health service facilities for all cases and controls (**Table S1**).

We adopted a case-control design with cases being defined as neighborhoods with one or more confirmed COVID-19 cases. Our level of analysis was neighborhood. One geocode was reported for each neighborhood regardless of the number of cases. For each case neighborhood, four controls were identified by determining the geocodes that were located 4500 meters (4.5 km) east, south, west and north of the COVID-19 (case) neighborhoods (see **Fig. S1** for a diagram illustrating this design). If there is another case community in 4.5 km from the control community, we selected another control community farther than 4.5 km from the two case communities. We calculated the numbers of each type of public facility in the circlular (“buffer”) areas surrounding the case and control neighborhoods. A radius of 1.5-km (approximately 1 mile) from the reported geocodes and the selected control neighborhood was used to capture information on nearby public facilities. Although there is no consensus on “proximity,” this cut-off point has been widely used in research (*14-16*).

A total of 986,363 POIs were gathered in 162 cities throughout China where public reporting of COVID-19 case locations was practiced. We excluded Wuhan city due to its exceptionally high incidence of COVID-19 and it is less possible to find the control neighborhood. We also excluded neighborhoods with no or few nearby public facilities (total number across eight types < 8) due to it may highly correlated with the lack of POI data recording lack of relevance of this study to these neighborhoods. We fit mutivariable logistic regression models adjusting for some city level covariates including population size, Gross Domestic Product (GDP), unemployment rate, Government Budget Balance, and population mobility recorded by the *Tencet* application (WeChat and QQ). Due to the high correlations between the eight types of facilities with Pearson's correlation coefficients ranging from 0.62 to 0.92 (**Table S2**), we ran separate multivariable models for each of the eight types. As we had eight variables of interest, we applied the Bonferroni method to control for multiple comparisons and used the *p*-value of <0.006 to indicate statistical significance (*18*).

Our sample included 4,329 case and 17,316 control neighborhoods. The case neighborhoods had 7,631 COVID-19 cases accounting for 23.0 % of the total number of cases outside of Wuhan City. The time (**Fig. S2**) and geographical distributions (**Fig. 1**) of the analyzed cases were consistent with the trends of the COVID-19 pandemic in China. Over 50% of publicly released COVID-19 cases were reported from Feb 1^st^ 2020 to Feb 10^th^ 2020 during the peak period of the outbreak. The provinces near the epicenter of Hubei Province had larger numbers of cases and higher proportions of cities reporting COVID-19 cases (**Fig. 1**) than other provinces.

**Figure 1.**
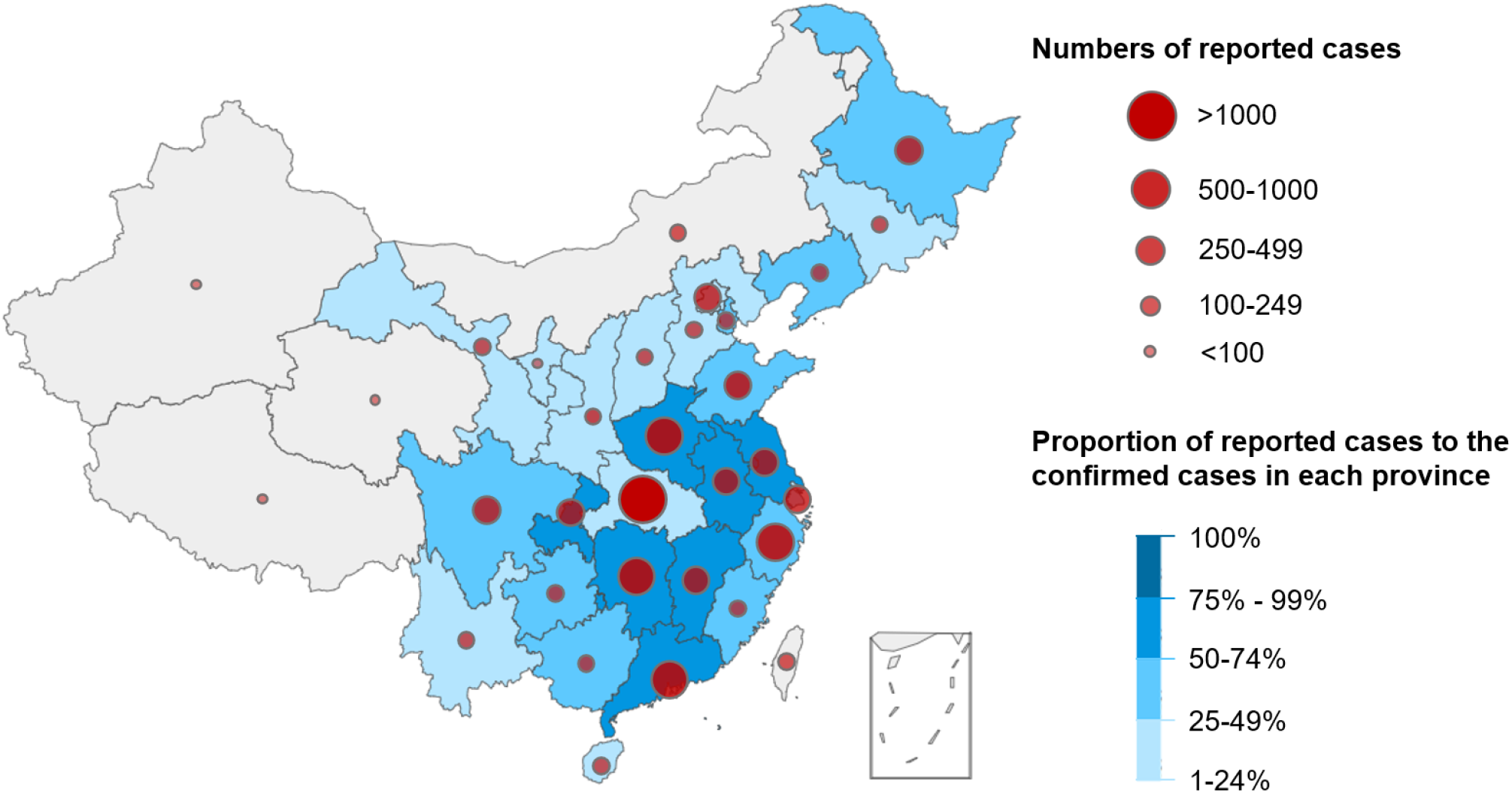
Geographic Distribution of Reported and Confirmed COVID-19 Cases in China.

Case neighborhoods had greater quantity of each of the eight types of public facilities compared to control neighborhoods (**Table 1**). However, the specific relationship between public facilities and reported cases of COVID-19 differed by facility type (**Fig. 2**). The relationship of six types of public facilities **—** shopping, restaurant, education, health service, hotel, and living facilities **—**and having COVID-19 cases in the community was nearly linear when the number of facilities was smaller than 10 and flattened thereafter (**Fig. 2, A,B, E-H**). The association between recreational facilities and having COVID-19 cases in the community was nearly linear (**Fig. 1, C**).The association of public transit with having COVID-19 cases in the community was J-shaped (**Fig. 2 D**).

**Figure 2.**
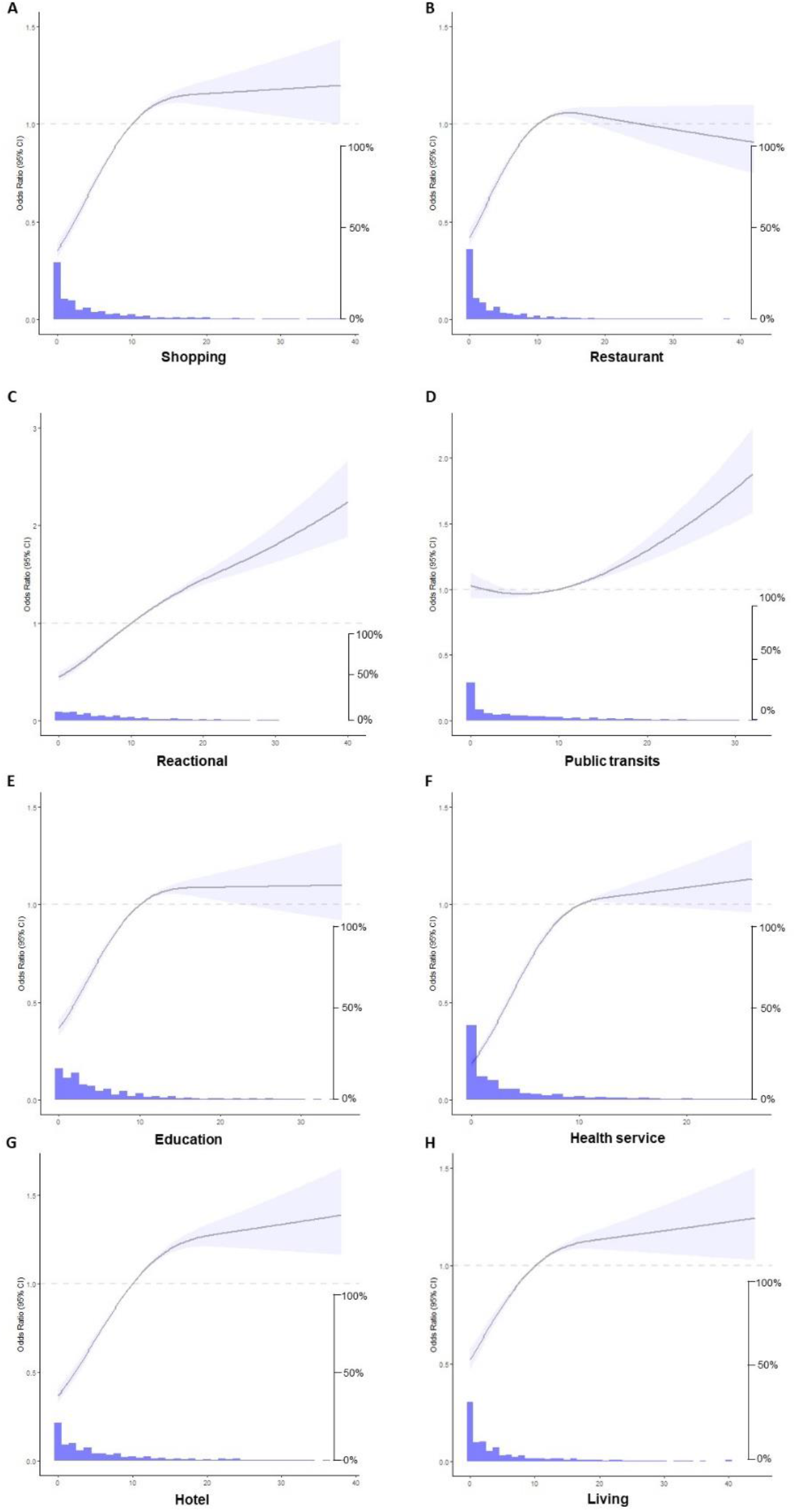
Association of eight types of public facilities with having COVID cases in the communities. Model adjusted for population size, Gross Domestic Product (GDP), unemployment rate, Government Budget Balance, average population mobility of each city from 18^th^ January 2017 to 30^th^ April collecting by *Tencent* applications (e.g. WeChat and QQ)

**Table 1.** Mean and median of the eight types of facilities.

| **Facilities** | **Control communities** | **COVID-19 communities** | **P-values\*** |
| --- | --- | --- | --- |
| **N** | **17,316** | **4,329** |  |
| **Restaurant** |  |  |  |
| Mean (SD) | 14.9 (42.2) | 17.6 (27.5) | <0.001 |
| Median (IQR) | 2.0 (0.0, 10.0) | 5.0 (1.0, 20.0) | <0.001 |
| **Shopping** |  |  |  |
| Mean (SD) | 14.9 (42.9) | 17.4 (24.7) | <0.001 |
| Median (IQR) | 3.0 (0.0, 10.0) | 7.0 (2.0, 21.0) | <0.001 |
| **Hotel** |  |  |  |
| Mean (SD) | 13.1 (26.9) | 19.4 (25.5) | <0.001 |
| Median (IQR) | 4.0 (1.0, 13.0) | 10.0 (3.0, 26.0) | <0.001 |
| **Living** |  |  |  |
| Mean (SD) | 14.7 (35.6) | 19.1 (28.0) | <0.001 |
| Median (IQR) | 3.0 (0.0, 12.0) | 6.0 (2.0, 25.0) | <0.001 |
| **Recreation** |  |  |  |
| Mean (SD) | 16.6 (29.6) | 23.9 (28.6) | <0.001 |
| Median (IQR) | 7.0 (2.0, 17.0) | 13.0 (5.0, 31.0) | <0.001 |
| **Public transits** |  |  |  |
| Mean (SD) | 2.0 (0.8) | 2.1 (0.8) | <0.001 |
| Median (IQR) | 2.0 (1.0, 3.0) | 2.0 (1.0, 3.0) | <0.001 |
| **Education** |  |  |  |
| Mean (SD) | 12.5 (26.5) | 15.7 (21.4) | <0.001 |
| Median (IQR) | 4.0 (1.0, 11.0) | 7.0 (3.0, 20.0) | <0.001 |
| **Health services** |  |  |  |
| Mean (SD) | 7.0 (17.2) | 19.3 (27.2) | <0.001 |
| Median (IQR) | 1.0 (0.0, 6.0) | 7.0 (2.0, 25.0) | <0.001 |
| **All facilities \*** |  |  |  |
| Mean (SD) | 104.9 (220.1) | 146.8 (169.7) | <0.001 |
| Median (IQR) | 34.0 (14.0, 92.0) | 68.0 (27.0, 206.0) | <0.001 |

\* Numbers shown are N (%) unless otherwise noted.
\* P-values was calculated by paired t-test and Mann–Whitney U test

In logistic regression models adjusting for city-level variables, having a larger number of facilities was associated with higher odds of community with COVID-19 cases (ORs ranged from 1.32 for public transits facility to 4.12 for health services; **Fig. 3**)

**Figure 3.**
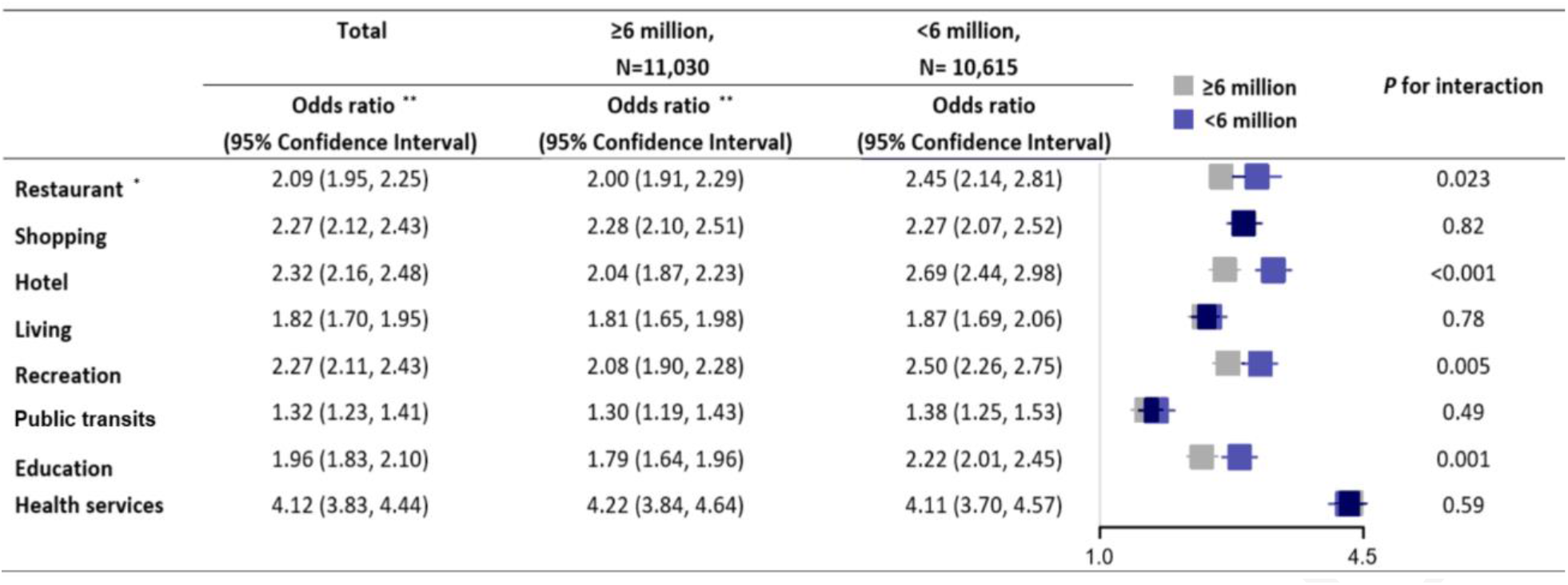
Sub-Group Analysis by City Population above and below 6 million: Odds Ratio and 95% CI of Having COVID-19 Cases in the Community with Eight Types of Surrounding Facilities. * The reference group was the “lower than the median” for each public facility ** Model adjusted for population size, Gross Domestic Product (GDP), unemployment rate, Government Budget Balance, average population mobility of each city from 18^th^ January 2017 to 30^th^ April collecting by *Tencent* applications (e.g. WeChat and QQ)

Next, we explored whether the association between each type of public facillity and having COVID-19 cases in the neighborhood differed by city's population size. We used a cut-off point of 6 million (representing approximately the median value in our sample) to classify larger versus smaller cities. The association between restaurants, hotels, and recreational and education facilities was more pronounced in smaller cities than larger ones (*P-values* for interaction <0.006, **Figure 3**).

Our study is novel and has several strengths. First, unlike previous studies only analyzing provincial or city-level data, our study utilized the addresses of reported COVID-19 cases linked with POI data to investigate the associations between neighborhood-level factors and COVID-19. Second, for neighborhoods that reported case locations, all confirmed cases were reported. Extensive testing and reporting of COVID-19 cases in China contributed to the reliability of our results. Third, control neighborhoods were systematically chosen to be compatible with case neighborhoods and our analyses also adjusted for city-level characteristics. More in-depth modelling with stratitication by city population size uncovered significant effect modification that may shed light on future disease containment strategies.

Our primary finding was that having COVID-19 cases in the neighborhood was associated with having larger numbers of neighborhood-level public facilities, particularly in cities with fewer than six million population. While this finding may seem intuitive, it has not been previously demonstrated. Empirical data-driven evidence from our study helps to address the controversies around more or less strict social distancing measures. One plausible explanation is that residents living in neighborhood with greater surrounding facilities are more attracted to go out and use these facilities, where there is greater exposure to the SARS-CoV-2 virus that increases the risk of infection. Also, in some facilities such as restaurants, hotels, recreation, education and health services, people are more likely to take off their masks to communicate or dine. In our subgroup analyses, we found that the associations of COVID-19 transmission with restaurants, hotels, recreation and education facilities were more prounouced in cities with population sizes smaller than six million compared to larger cities. One potential explanation for the modification effect of population size was that the geographical scopes of the activities of asymptomatic individuals in larger cities were wider and more disperse due to its relave convenient transportation; thus infection may occur far from its residential neighborhood, which was beyond the scope of our study. The tracking of cases in Beijing, China in June 2020 partially confirmed this hypothesis. Another explanation is that in smaller cities, the social distancing polices may be not implemented as well as in larger cities.

Our study has some implication for the policy makers to design the targeted intervention strategy. To be more specific, in the neighborhood with more public facilities surrouding, more extensive preventative measures such as educational or behavioral enhancements for mask-wearing and social distancing are needed. Additionally, those facilities nearby also needs to be addressed on how to **MANAGE** them. For example, partially shut down or restricted hours, staggered appointments/reservations, different hours for different subpopulations of the neighborhood, specific reminder for social distancing in those facilities may be useful for prevention during the pandamic. With those targeted intervention, policy makers and social forces can optimize resource allocations in those settings with limited medical and personnel resources.

Our findings are relevant for COVID-19 control in the long term because it sheds lights on neighborhood-level factors associated with transmission. Up to now, there is still no effective vaccine or drug treatments for COVID-19, and the primary intervention is non-pharmaceutical and mostly preventive through public health approaches. A recent modelling studies indicated that without the non-pharmaceutical interventions in China, COVID-19 cases in China would likely have shown a 67-fold increase by February 29, 2020 (*19*). Another study found that forty percent of asymptomatic cases became seronegative and 12.9% of symptomatic patients tested negative for SARS-CoV-2-specific immunoglobulin G antibodies two or three months after the infection (*20*). Therefore, active surveillance and social distancing may be needed for longer periods than previously anticipated, which might pose substantial social and economic burdens. Hence, neighborhood-based facility-targeted prevention at relevantly low costs to hinder COVID-19 transmission is desired.

Different countries and even different regions within one country have adopted various disease control strategies with vastly different results and socio-economic consequences. Our study findings have implications for other low-and middle-income countries such as India and Brazil with similar population density and city infrastructures but limited human and economic resources to fight the surging epidemic. If validated in other countries and by further research, targeted prevention strategies by city size is warranted and may lead to better disease control through facility-based containment approaches.

Our study has limitations. First, we did not have location data for all cases in China. Nevertheless, we collected data on all publicly reported cases for four months covering the major epidemic period in China. Second, we did not have detailed information on routes of COVID-19 transmission. Our cross-sectional study can not make causal inferences regarding the relationship between neighborhood features and disease transmission. Third, the POI data is not extensive recorded in some region with lower local economic status and we have excluded those cases neighborhood with total POIs lower than eight. More studies are needed, especially epidemiological case tracking data with extensively recorded geo-information and longitudinal research.

In summary, having COVID-19 cases in residential neighborhoods was associated with the numbers and many types of surrounding public facilities. The associations of COVID-19 transmission with restraurants, hotels, recreation and education facilities were more pronounced in Chinese cities with a population size fewer than six million. COVID-19 has caused millions of cases and claimed hundreds of thousands of lives, and it is very likely that our battle against it will not be over soon. Large-scale disease control strategies such as social distancing and lock-downs have been shown to be effective, but achieved at enormous social and economic costs. Targeted interventions with lower costs and high efficiencies are warranted. We expect our findings to shed light on improving the COVID-19 prevention strategies at the neighborhood level for China and potentially other countries.

## Data Availability

The data, analytical methods, and study materials will be made available to other researchers for purposes of reproducing the results or replicating the procedure from the corresponding author (X.J.) on reasonable request.

## ACKNOWLEDGEMENTS

We thank thousands of health professionals, community workers, and Centers for Disease Control staff in China who collected data and continue to work to contain COVID-19 in China and elsewhere.

## Funding

This study is not funded by any external sources. No funder had any role in study design, data collection and analysis, the decision to publish, or in preparation of the manuscript.

## Author contributions

X.J., L.L.Y., and C.W. designed the study. X.J. and Y.L. collected and processed the API and POI data. X.J. and Y.L. conducted the analyses. X.J. and Y.L. wrote the manuscript. E.G., S.X., Y.Y., K.C., R.V., L.L.Y. and C.W. assisted with interpretation of the results and edited the manuscript. All authors critically reviewed and approved the manuscript.

## Competing interests

All authors declare no competing interests.

## Notes

### Competing Interest Statement

The authors have declared no competing interest.

## REFERENCES

1. N. Zhu et al., A Novel Coronavirus from Patients with Pneumonia in China, 2019. N Engl J Med 382, 727–733 (2020).

2. N. Lu, K. W. Cheng, N. Qamar, K. C. Huang, J. A. Johnson, Weathering COVID-19 storm: Successful control measures of five Asian countries. Am J Infect Control 48, 851–852 (2020).

3. J. Xie, Y. Zhu, Association between ambient temperature and COVID-19 infection in 122 cities from China. Sci Total Environ 724, 138201 (2020).

4. H. Lau et al., The association between international and domestic air traffic and the coronavirus (COVID-19) outbreak. J Microbiol Immunol Infect 53, 467–472 (2020).

5. N. Sattar, I. B. McInnes, J. J. V. McMurray, Obesity Is a Risk Factor for Severe COVID-19 Infection: Multiple Potential Mechanisms. Circulation 142, 4–6 (2020).

6. J. B. Dowd et al., Demographic science aids in understanding the spread and fatality rates of COVID-19. Proc Natl Acad Sci U S A 117, 9696–9698 (2020).

7. J. W. Tang, Y. Li, I. Eames, P. K. Chan, G. L. Ridgway, Factors involved in the aerosol transmission of infection and control of ventilation in healthcare premises. J Hosp Infect 64, 100–114 (2006).

8. M. Jayaweera, H. Perera, B. Gunawardana, J. Manatunge, Transmission of COVID-19 virus by droplets and aerosols: A critical review on the unresolved dichotomy. Environ Res 188, 109819 (2020).

9. L. Debenham, J. Reynolds, Climbing Gyms as Possible High-Risk Transmission Locations in Microbial Outbreaks. Wilderness Environ Med, (2020).

10. M. Stromgren et al., Place-based social contact and mixing: a typology of generic meeting places of relevance for infectious disease transmission. Epidemiol Infect 145, 2582–2593 (2017).

11. M. Winters et al., Where do they go and how do they get there? Older adults’ travel behaviour in a highly walkable environment. Soc Sci Med 133, 304–312 (2015).

12. R. Shigematsu et al., Age differences in the relation of perceived neighborhood environment to walking. Med Sci Sports Exerc 41, 314-321 (2009).

13. U. N. Emeruwa et al., Associations Between Built Environment, Neighborhood Socioeconomic Status, and SARS-CoV-2 Infection Among Pregnant Women in New York City. JAMA, (2020).

14. C. Coutts, T. Chapin, M. Horner, C. Taylor, County-level effects of green space access on physical activity. J Phys Act Health 10, 232–240 (2013).

15. Y. K. Ranchod, A. V. Diez Roux, K. R. Evenson, B. N. Sanchez, K. Moore, Longitudinal associations between neighborhood recreational facilities and change in recreational physical activity in the multi-ethnic study of atherosclerosis, 2000-2007. Am J Epidemiol 179, 335–343 (2014).

16. A. T. Kaczynski et al., Are park proximity and park features related to park use and park-based physical activity among adults? Variations by multiple socio-demographic characteristics. Int J Behav Nutr Phys Act 11, 146 (2014).

17. H. Wang, M.-P. Kwan, M. Hu, Social exclusion and accessibility among low- and non-low-income groups: A case study of Nanjing, China. Cities 101, 102684 (2020).

18. A. C. Leon, Multiplicity-adjusted sample size requirements: a strategy to maintain statistical power with Bonferroni adjustments. J Clin Psychiatry 65, 1511–1514 (2004).

19. S. Lai et al., Effect of non-pharmaceutical interventions to contain COVID-19 in China. Nature, (2020).

20. Q. X. Long et al., Clinical and immunological assessment of asymptomatic SARS-CoV-2 infections. Nat Med 26, 1200–1204 (2020).

